# Combined PD-L1 and TIM-3 blockade improves the expansion of fit human CD8+ antigen-specific T cells for adoptive immunotherapy

**DOI:** 10.1101/2021.08.30.21262835

**Authors:** Shirin Lak, Valérie Janelle, Anissa Djedid, Gabrielle Boudreau, Ann Brasey, Véronique Lisi, Cédric Carli, Lambert Busque, Vincent-Philippe Lavallée, Jean-Sébastien Delisle

**Affiliations:** Centre de recherche de l’Hôpital Maisonneuve-Rosemont, Montréal, Qc, Canada; Centre de recherche du CHU Sainte-Justine, Montréal, Qc, Canada; Biomarker Unit, Centre C3i, Montréal, Qc, Canada; Department of Medicine, Université de Montréal, Montréal, Qc, Canada; Hematology-Oncology and Cell therapy Division, Hôpital Maisonneuve-Rosemont, Montréal, Qc, Canada; Department of Pediatrics, Université de Montréal, Montréal, Qc, Canada; Hematology-Oncology Division, CHU Sainte-Justine CHU Sainte-Justine

## Abstract

**Background:** The stimulation and expansion of antigen-specific T cells *ex vivo* enables the targeting of a multitude of cancer antigens. However, clinical scale T-cell expansion from rare precursors requires repeated stimulations *ex vivo* leading to T-cell terminal effector differentiation and exhaustion that adversely impact therapeutic potential. We leveraged immune checkpoint blockade relevant to antigen-specific CD8+ human T cells to improve the expansion and function of T cells targeting clinically relevant antigens.

**Methods:** A clinically-compliant protocol relying on peptide-pulsed monocyte-derived dendritic cells and cytokines was used to expand antigen-specific CD8^+^ targeting the oncogenic Epstein-Barr virus (EBV) and the tumor associated antigen (TAA) Wilms Tumor 1 (WT1) protein. The effects of antibody-mediated blockade of immune checkpoints applied to the cultures (T-cell expansion, phenotypes and function) were assessed at various time points. Genomic studies including single cell RNA (scRNA) sequencing and T-cell receptor sequencing were performed on EBV-specific T cells to inform about the impact of immune checkpoint blockade on the clonal distribution and gene expression of the expanded T cells.

**Results:** Several immune checkpoints were expressed early by *ex vivo* expanded antigen-specific CD8^+^ T cells, including PD-1 and TIM-3 with co-expression matching evidence of T-cell dysfunction as the cultures progressed. The introduction of anti-PD-L1 (expressed by the dendritic cells) and anti-TIM-3 antibodies in combination (but not individually) to the culture led to markedly improved antigen-specific T-cell expansion based on cell counts, fluorescent multimer staining and functional tests. This was not associated with evidence of T-cell dysfunction when compared to T cells expanded without immune checkpoint blockade. Genomics studies largely confirmed these results, showing that double blockade does not impart specific transcriptional programs or patterns on TCR repertoires. However, our data indicate that combined blockade may nonetheless alter gene expression in a minority of clonotypes and have donor-specific impacts.

**Conclusions:** The manufacturing of antigen-specific CD8^+^ T cells can be improved in terms of yield and functionality using blockade of TIM-3 and the PD-L1/PD-1 axis in combination. Overcoming the deleterious effects of multiple antigenic stimulations through PD-L1/TIM-3 blockade is a readily applicable approach for several adoptive-immunotherapy strategies.

## Background

The vast majority of potentially actionable cancer antigens are major histocompatibility (MHC)-bound peptides. These include tumor-specific antigens (TSA) resulting from mutated proteins or aberrantly expressed regions of the genome, tumor associated antigens (TAA) that come from abnormally expressed self-proteins and finally, non-self peptides best described in the setting of virus-associated cancers or allogeneic stem cell transplantation^1 2^. While it has been possible to isolate TSA or TAA-specific T-cell receptors (TCR) and manufacture TCR-transgenic T cells *ex vivo* followed by their injection in patients, such adoptive immunotherapy approaches face numerous technical hurdles and are currently available only for a minority of relevant cancer antigens^3^. Several effective strategies rely on the *ex vivo* expansion of native antigen-specific T cells, enabling the targeting of a vast array of antigens. However, the expansion of large numbers of antigen-specific T cells requires repeated antigen exposure (through co-culture with antigen-presenting cells – APC) and stimulatory cytokines, potentially leading to T-cell dysfunction (terminal effector differentiation and exhaustion) and poor performance following adoptive transfer^4-6^.

Alternatively, the endogenous cancer-reactive T-cell repertoire can be mobilized through the systemic administration of antibodies that prevent signaling from inhibitory co-signaling receptors present on the surface of exhausted T cells^7 8^. Such “immune-checkpoint” blockade, targeting most commonly Cytotoxic T-lymphocyte-associated protein 4 (CTLA-4) and Programmed death-1 (PD-1) on T cells (or its corresponding primary ligand, Programmed death-ligand 1 - PD-L1), is now the cornerstone of therapeutic regimes against several types of neoplasia including advanced melanoma and lung cancer^9^. As dysfunctional cancer-reactive T cells often express multiple negative co-signaling molecules, a strategy has been to use combined approaches with the caveat that increased response may come with more immune-related toxicities^10^. *Ex vivo* expanded T cells express inhibitory receptors, and PD-L1/PD-L2 silenced antigen-presenting dendritic cells have been shown to improve the expansion and function of antigen-specific T cells *ex vivo*^11^, thus providing a solid rationale to leverage immune checkpoint blockade to improve T-cell manufacturing for adoptive immunotherapy.

We show herein that the *ex vivo* expansion of CD8+ T cells specific for an oncogenic virus antigen and a TAA is enhanced by using the combined blockade of PD-L1 and the immune checkpoint T-cell immunoglobulin and mucin-containing protein-3 (TIM-3), while single blockade of either receptor failed to improve T-cell expansion. Increased antigen-specific T-cell expansion under dual immune checkpoint blockade was not associated with phenotypic and functional evidence of exhaustion or terminal effector differentiation. This was corroborated with single-cell RNA (sc-RNA) and VD(J) sequencing, which nonetheless revealed that dual immune checkpoint blockade may impact T-cell gene expression in a donor and clonotype-dependent fashion. Taken together, our results show that dual PD-L1/TIM-3 blockade during *ex vivo* expansion can yield large quantities of fit human antigen-specific T cells for adoptive immunotherapy.

## Methods

### Donors and cellular procurement

Peripheral blood mononuclear cells (PBMCs) from HLA-A0201 expressing volunteer donors were isolated using Ficoll-Hypaque gradient (STEMCELL Technologies) from fresh whole blood (collected by venipuncture) or leukoreduction system chambers provided by Héma-Québec (LRSCs) as previously described^12 13^. All donors provided written informed consent, and all the experiments were approved by the Héma-Québec and Hôpital Maisonneuve-Rosemont Research Ethics Committees. Recovered PBMCs were either used immediately for experiments or cryopreserved for future use in 90% fetal bovine serum and 10% dimethyl sulfoxide (DMSO).

### Dendritic Cells (DCs) differentiation and antigen pulsing

Monocyte isolation and DC differentiation have been generated as previously described^4^. Briefly, monocytes were obtained using the adherence method whereby PBMCs were plated in adherent plastic plates (Sarstedt) in media (X-Vivo 15 medium (LONZA) supplemented with 5% human serum, 2 mM L-glutamine and 1mM Sodium Pyruvate (Gibco), 1000 U/ml (100 ng/ml) IL-4 and 800 U/ml (50ng/ml) GM-CSF (Both from STEMCELL technologies)) and incubated in a CO_2_, 37°C incubator for 7 days. On day 4, media was replaced with fresh media supplemented with IL-4 and GM-CSF. On day 7, DCs were matured with maturation media containing 1000 U/ml (100 ng/ml) IL-4, 800 U/ml (50ng/ml) GM-CSF (Both from STEMCELL technologies), 10 ng/ml TNF-α (STEMCELL technologies), 1 μg/ml PGE2 (SIGMA), 10 ng/ml IL-1β (Feldan), 100 ng/ml IL-6 (Miltenyi Biotec) and loaded with desired peptide (1 μg/mL Latent membrane protein 2 - LMP2_426-434_ (CLGGLLTMV) or 1 μg/mL Wilm’s Tumor 1 - WT1_37-45_ (VLDFAPPGA), both from JPT Peptides. Lastly, DC media was supplemented with IFN-γ 1000U/ml (Feldan) for the last 24 hours of maturation.

#### *Ex Vivo* expansion of antigen-specific T cells

Antigen-specific T cells were stimulated using a clinically compliant protocol as previously described (^4^ and www.clinicaltrials.gov NCT03091933) from 15×10^6^ PBMCs and expanded through multiple weekly stimulations using irradiated (40 Gy) autologous, peptide-loaded monocyte-derived DCs at a 1:10 (DC:PBMC) in a G-Rex6® Well Plate vessel (Wilson Wolf Manufacturing, New Brighton, MN). Our complete T-lymphocyte culture (CTL) media (Advanced RPMI 1640, 10% human serum, 1X L-glutamine (Gibco)) was supplemented with the following cytokines; week 1: IL-21 (30 ng/mL) and IL-12 (10 ng/ mL) (Both from Feldan), week 2: IL-21, IL-2 (100 IU/mL), IL-7 (10 ng/mL) and IL-15 (5 ng/mL) (STEMCELL Technologies), subsequent weeks: IL-2, IL-7 and IL-15. Medium, including cytokines, was refreshed every 3 to 4 days, and antigen re-stimulation was done every week with peptide-loaded monocyte-derived DCs. Cell concentration was adjusted to 1:10 ratio each week. When indicated, cultures were supplemented with 20 μg/ml of anti-PD-L1 blocking mAb (BioXcell, 29E.2A3) or/and 10 μg/ml of anti-TIM3 blocking mAb (Biolegend, F38-2E2) with all media changes. Respective isotype antibodies, mouse IgG1 (BioXcell, MOPC-21) and InVivoMAb mouse IgG2b (BioXcell, MPC-11), were used at the same concentration where indicated. All cell cultures were performed in monitored incubators (37°C in 5% CO_2_ and 5% air humidity). Cell viability and cell counts were assessed by the countess automated cell counter (Invitrogen) using trypan blue (Invitrogen) at a 1:1 ratio with the cellular suspension in cell counting chamber slides (Invitrogen-C10283).

### Flow cytometry

The phenotype of mature DCs was assessed by cell surface expression of the following markers (antibody clone in parenthesis) CD80 (L307.4), CD86 (2331, FUN1), HLA-ABC (W6-32), CD11c (3.9), CD19 (HIB19) (all from BD Biosciences)), HLA-DR (LN3), and CD83 (HB15e) (both from Invitrogen). To detect antigen-specific CD8^+^ T cells, up to 10^6^ cells were suspended in phosphate-buffered saline (PBS) plus 2% fetal bovine serum (FBS) and stained with appropriate allophycocyanin (APC) labeled MHC-Dextramers A*0201 (Immudex,), for 10 minutes in the dark at 4°C. For additional cell surface markers and phenotyping, cells were stained with following antibodies: CD3 (SKY7), CD3 (UCHT1), CD8 (SK1), CD45RO (UCHL1), CD45RA (5H9), CCR7 (150503), LAG3 (T47.530), and CD4 (RPA-T4) all from BD Biosciences; CD62L (DREG-56), TIM3 (F38-2E2), PD-1 (EH12.2H7), KLRG1 (2F1/KLRG1), CD57 (HCD57), and PD-L1 (29E.2A3) from Biolegend and CD8 (RPA-T8) and CD244 (eBioDM244) from Invitrogen. Staining was performed at room temperature (RT) in the dark for 30 minutes. Intracellular staining was done with the Foxp3 / Transcription Factor Staining Buffer Set as recommended by the manufacturer (eBioscience). Before the fixation step, up to 10^6^ cells were stimulated to produce cytokines with the indicated peptide (0.5 μg/ml) (test condition) or PMA (50 ng/mL)-ionomycin (500 ng/mL) (Sigma-Aldrich) (positive control) and a pool of irrelevant peptides (from the cytomegalovirus pp65 protein - PepMix™ HCMVA, JPT peptides) for 4 h at 37°C. Cells are suspended in CTL media plus brefeldin A (Biolegend) to block the secretion of cytokines during the stimulation period. Cells were then harvested and stained for cell-surface markers, including CD3, CD4, and CD8 in 4 °C for 20 minutes. Next, fixed and permeabilized cells were stained with intracellular cytokine detection antibodies; IFN-γ (4S.B3), IL-2 (MQ1-17H12), and TNF-α (Mab11) all purchased from BD Biosciences) and Ki67 (ki-67) from Biolegend at RT for 20 minutes. Cell acquisition was performed on a Fortessa or LSR II flow cytometer (BD Biosciences), and data were analyzed with Flowlogic Software (inivai).

### IFN-γ Enzyme-linked immune-spot (ELISpot)

ELISPOT assays were performed using Human IFN-ELISpot^PLUS^ kit (MABTECH), following the manufacturer’s instruction. Cultured cells were added (5×10^4^) to wells in duplicates and then stimulated with anti-CD3 mAb (positive control), irrelevant peptides (specificity control), test peptide or vehicle only (DMSO). Spots were counted using ELISpot reader (vSpot Reader Spectrum, AID).

### Flow Cytometry-based Cytotoxicity assay

Flow cytometry-based cytotoxicity assay was performed using either Cell Trace Violet (CTV) or Cell Trace Yellow (CTY) (Invitrogen Life Technologies) labeled LMP2_426-434_ pulsed as target cells as described before^14^. Briefly, target cells were prepared by stimulating autologous PBMCs with the T-cell mitogen phytohemagglutinin (PHA) (3 × 10^6^ PBMCs/mL were incubated in T-cell media with 20 μg/mL PHA) for 3 days at 37 °C and 5% CO_2_. Peptide pulsed (or not pulsed) target cells were co-cultured with LMP2-specific T cells at various ratios for 4 hours in CTL plus 10% horse serum. Unpulsed target cells alone were used as control. After incubation, cells were harvested, stained with LIVE/DEAD Fixable Aqua Dead Cell Stain Kit, Life Technologies) according to manufacturers’ instructions and viable target, cells were quantified by flow cytometry using Flow Count Beads (Beckman Coulter). Cytotoxicity was calculated by comparing the percentage of viable target cells in test conditions relative to control (100 - (Target cell alive/ Target cell alone) × 100).

### Single-cell RNA sequencing and High-throughput TCR sequencing

At indicated time points, T cells were sorted based on MHC-Dextramer and CD8 staining using FACS Aria III (DB Biosciences). Post sorted purity of the sorted population was 94%. A fraction of the sorted antigen-specific T cells was subjected to both VDJ and transcriptome sc-RNA sequencing at the Genome Quebec facilities. Briefly, cells were counted and viability assessed using a hemocytometer and Trypan Blue. The targeted cell recovery was set at 6000 cells and libraries were prepared using the kits Chromium Next GEM Single Cell 5’ Kit v2 and Chromium Single Cell Human TCR Amplification Kit (10x Genomics inc.), as per the manufacturer’s recommendations. Libraries were quantified using the Kapa Illumina GA with Revised Primers-SYBR Fast Universal kit (Kapa Biosystems). Average size fragment was determined using a LabChip GX (PerkinElmer) instrument. The 10X Single cells 5’ libraries were sequenced on Illumina HiSeq 4000 PE28×98 while the 10X Single Cells VDJ (Human T) libraries were sequenced on Illumina HiSeq 4000 PE150. The Illumina control software was HCS HD 3.4.0.38, the real-time analysis program was RTA v. 2.7.7. bcl2fastq2 v2.20 was then used to demultiplex samples and generate fastq files. Reads were aligned to GRCh38 genome assembly, gene expression matrices generated, and clonotype identification performed using CellRanger v3.0.2. The resulting gene expression matrices were normalised by total UMI counts per cell multiplied by the median UMI count per cell and natural-log transformed using Scanpy v1.4^15^. PhenoGraph^16^ clustering was applied with k=20 and clusters with a median mitochondrial fraction greater than 0.2 were filtered out. Clonotypes were defined by nucleotide sequences of alpha and beta chains. In order to remove potential doublets, we restricted our analysis to cells in which exactly one alpha and one beta chain were identified. Furthermore, we retained only the most abundant clonotype for each alpha and beta chain. The number of retained cells per sample is provided in Table S1. After filtering, individual count matrices were combined and normalized and log transformed, as described above, for downstream analyses. Pathway expression is defined as the log2 of the sum of the normalized gene expression of individual genes. Genes included with each pathway are listed in Table S2 (taken from^17^). Significance of the changes in pathway expression between the control (CTRL) and delayed double blockade (DDB) conditions was assessed using a Mann-Whitney U test. To set the threshold at which p-values are considered significant, we compared the p-values obtained with the pathway of interest to those obtained with random sets of genes. For each pathway of interest, we generated 1000 random gene sets with the same number of genes drawn from the same expression distribution. On each of these random gene sets, we performed the CTRL vs DDB comparison and recorded the p-values obtained from the Mann-Whitney U test and report the percentile ranking of the p-value in the pathway compared to the p-values obtained from the random gene sets.

Bulk TCR repertoire profiling was performed from sorted multimer positive and negative CD8^+^ T-cell RNA preserved in Trizol reagent (Invitrogen) using Next-Generation Sequencing (NGS) targeting the hypervariable complementarity-determining region 3 (CDR3) of the T cell receptor beta chain (TCRβ). TRIzolTM extracted RNA in combination with the PureLink RNA mini and microcolumn system (Thermo Fisher), quantified by UV spectrophotometry (Tecan), and QC was completed on Bioanalyzer chip (Agilent). TCRb amplicon libraries were prepared from 25 ng total RNA with the Oncomine™ TCR Beta-SR Assay for RNA (ThermoFisher Scientific). The TCRb libraries obtained were quantified on the ViiA 7 Real-Time PCR System with the Ion Library Taqman Quantitation kit (Thermo Fisher). NGS was completed on the Ion S5 semiconductor platform using an Ion 540 chip, prepared with the Ion Chef System (all from Thermo Fisher). TCRβ repertoire analysis was completed using the Ion Reporter Software (Thermo Fisher) and Immunarch package^18^ analyses using R software. The data for all genomic analyses can be found in the Gene Expression Omnibus (GSE182537 and GSE181682).

### Statistical Analysis

Statistical significance was analyzed were with the R software, version 4.0.4. Multiple group comparisons were performed using one-way ANOVA and Turkey post hoc test or the Kruskal-Wallis test with the Holm procedure to correct for multiple testing (if ANOVA requirements were not met). Paired Wilcoxon-Mann-Whitney tests were performed for two-group comparisons. p values of 0.05 or less were considered significant.

## Results

### Multiple stimulations are detrimental to antigen-specific T-cell expansion *ex vivo*

Antigen-specific T cells were stimulated using moDCs loaded with the peptide LMP2_426-434_ (CLGGLLTMV), which is an HLA-A0201 restricted antigen from the oncogenic Epstein-Barr virus (EBV). Weekly *ex vivo* stimulations were performed in cytokine supplemented media, and antigen-specific expansion was assessed before each stimulation. While overall T-cell expansion progressed following each of four stimulations (albeit at limited pace after day 14 and two stimulations), the percentage and absolute number of antigen-specific CD8 T cells, as identified by fluorescent HLA-0201/ LMP2_426-434_ multimer, stagnated despite additional stimulations (Figure 1A-C). Further, this halted growth was matched with a gradual change in phenotype. The predominance of central memory (Tcm) differentiation profile at day 14 evolved towards an effector memory (Tem) and effector (Teff) differentiation phenotype on days 21 and 28 (Figure 1D). These results were anticipated as serial T-cell stimulations have been associated with the development of effector T cells that gradually lose their capacity to expand and persist after adoptive transfer^6^. The expression of the inhibitory receptors related to T-cell exhaustion (PD-1, TIM-3, and to a lesser extent LAG3 and 2B4) was substantial on a significant fraction of antigen-specific T cells at day 14, with little modulation over time in culture (Figure 1E). However, the fraction of cells showing dual expression of PD-1 and TIM-3 increased with repeated stimulation (Figure 1F) in line with the development of CD8+ T-cell exhaustion^19-21^. Phenotyping of the moDCs revealed prevalent expression of corresponding PD-1 and TIM-3 ligands, (respectively PD-L1 and CEACAM-1/Galectin 9) (Figure 1G). These results confirm and extend previous data, suggesting that the early and persistent expression of PD-1 and TIM-3 by CD8^+^ T cells, along with the expression of their ligands by stimulating DCs, may represent a significant hurdle for the expansion of antigen-specific T cells for immunotherapy^19 20^.

**Figure 1.**
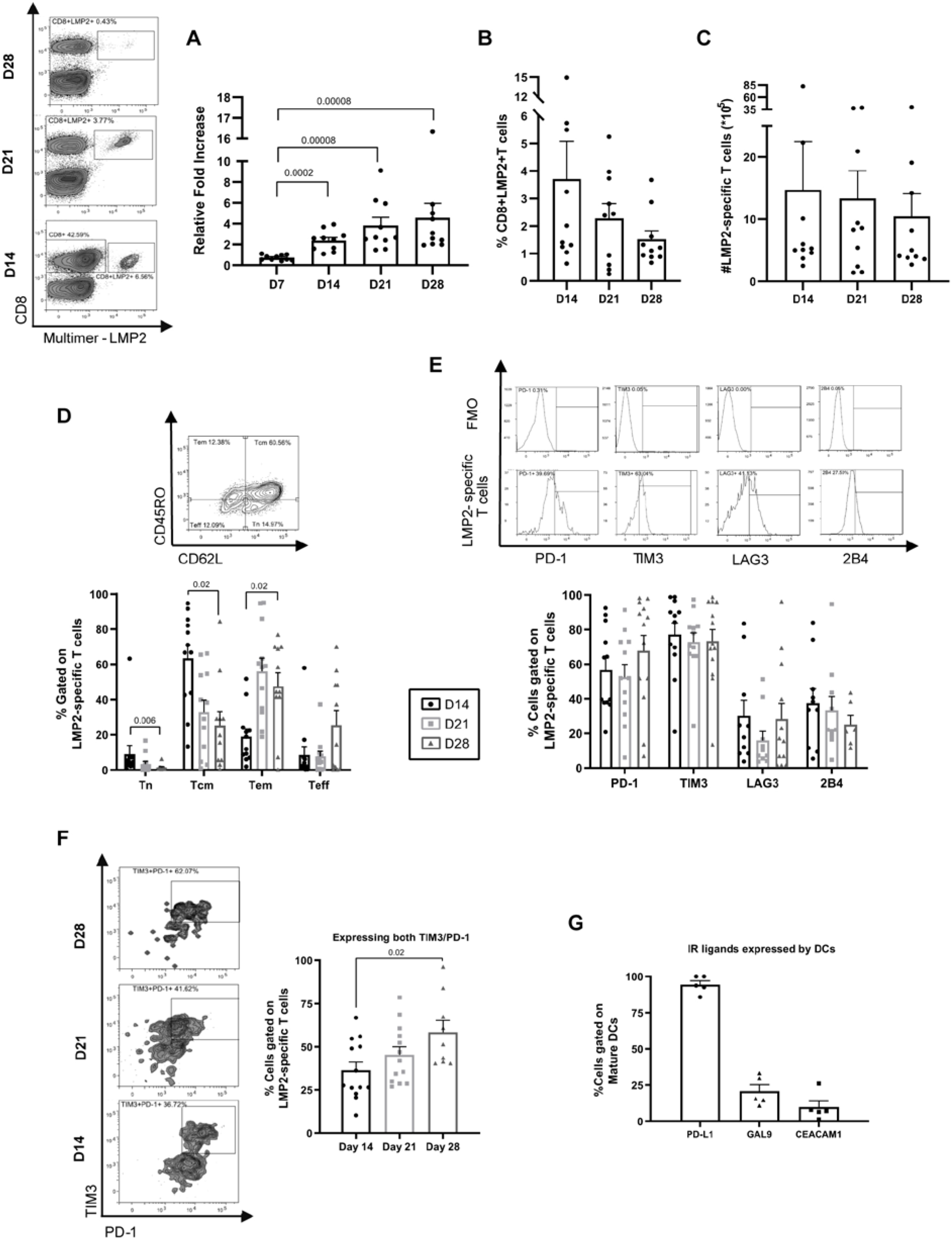
Repeated antigenic encounters ex vivo leads to antigen-specific T-cell exhaustion. (A) Representative fluorescent multimer staining of LMP2_426-434_-specific CD8^+^ T cells (LMP2) and total cellular expansion (expressed as fold change relative to culture input – 15×10^6^ cells). (B) Percentages, as well as (C) calculated number of antigen-specific T cells at the indicated time points (n=10 different donors). (D) Representative staining to determine naïve (Tn), central memory (Tcm), effector memory (Tem), and effector T cells (Teff) and the percentage of HLA-A02/LMP2_426-434_ multimer positive CD8^+^ T cells within these categories at different culture time points (n= 12 different donors). (E) Representative histograms of immune checkpoint expression evaluated relative to fluorescence minus one (FMO) control in HLA-A0201-LMP2_426-434_ (LMP2) multimer-stained T cells and compiled results from 12 different donors. (F) Representative staining of PD-1 and TIM-3 co-expression and compiled results from 9-13 different donors. (H) Percentage expression of PD-1 and TIM-3 ligands on dendritic cells used to stimulate T cells (n=5 different donors). Significant p values are indicated on figure panels. All error bars represent standard deviation to the mean (SEM).

### Combination of PD-L1/PD-1 axis and TIM-3 blockade significantly increases antigen-specific CD8^**+**^ T-cell expansion

In order to assess if immune checkpoint blockade during *ex vivo* expansion may improve antigen-specific T-cell yield, anti-PD-L1, anti-TIM-3, or both were added to the culture media at the beginning of culture and with all media changes. Cell counts at day 14, and even more strikingly at day 21, revealed that the double blockade condition significantly increased T-cell expansion relative to control (no checkpoint blockade) and the single blockade groups (Figure 2A). This translated into a marked increase in LMP2_426-434_-specific T-cell yield in the dual blockade condition, most evident at day 21 of culture (Figure 2B-C). In contrast, the single blockade of PD-L1 or TIM-3 offered no advantage at any time point. The phenotypic assessment of LMP2_426-434_-specific T cells revealed no statistically significant difference in the percentages of Tcm, Tem, and Teff and no difference in PD-1 or TIM-3 expression (Figure 2D-E). However, the percentage of T cells expressing the inhibitory receptors LAG3 and 2B4 was lower in the combined relative to the control condition. Hence, dual PD-L1 and TIM-3 blockade increased T-cell growth without conferring phenotypic changes associated with increased T-cell dysfunction.

**Figure 2.**
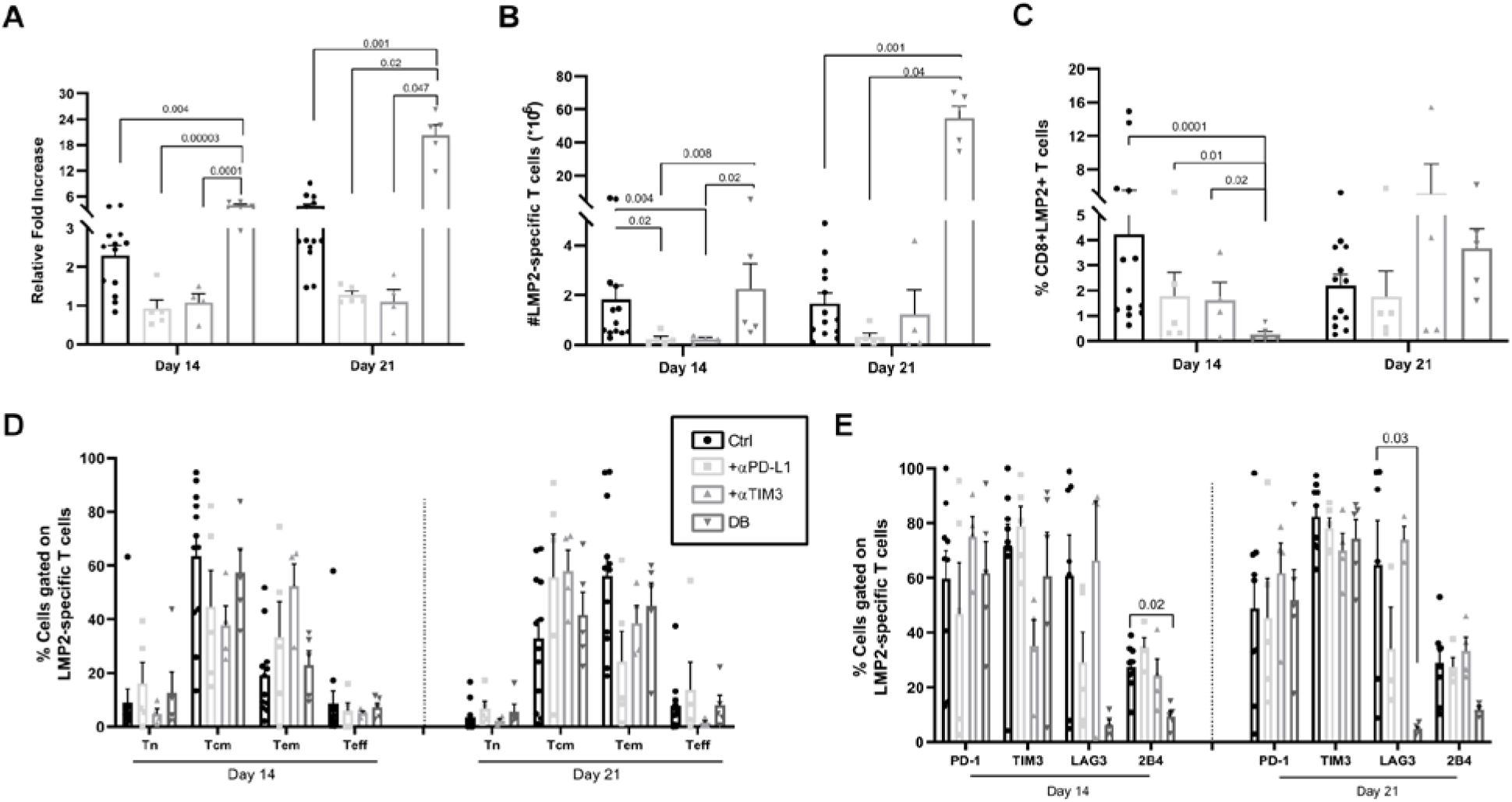
Dual but not single PD-L1 and TIM-3 blockade improves T-cell expansion. (A) Total cell expansion relative to input at the beginning of the culture in function of time and culture condition; no blocking antibodies (ctrl), anti-PD-L1 (α-PD-L1), anti-TIM3 (α-TIM3) or both (double blockade - DB). Absolute count (B) and percentage (C) of HLA-A0201-LMP2_426-434_ (LMP2) multimer positive T cells in the same conditions and at the same time-points. (D) T-cell differentiation phenotypes; naïve (Tn), central memory (Tcm), effector memory (Tem) and effector T cells (Teff) and (E) percentages of LMP2 multimer positive T cells expressing immune checkpoints in the function of culture conditions and time-points (n=4-10 different donors). Significant (<0.05) p-values are indicated on the figure panels. All error bars represent SEM.

It is known that TIM-3 has a dual function. TIM-3 is transiently upregulated at intermediate levels on activated T cells and confers activation signals^22^. However, in settings of chronic stimulation in the presence of its ligands, TIM-3 inhibits T-cell activation and behaves as a *bona fide* immune checkpoint. We therefore, slightly modified our protocol to introduce TIM-3 blockade at day 7, a week after the first stimulation (hereafter designated as delayed double blockade-DDB), with the expectation that it would further improve antigen-specific T-cell yield. Compared to the dual blockade started on day 0, DDB slightly increased total T-cell expansion at day 21 and more convincingly following an additional antigenic stimulation (day 28) (Figure 3A). Moreover, the DDB approach increased the percentage of LMP2_426-434_-specific T cells at all time points relative to the control condition (no blocking antibodies used), including at day 14 which was not the case when PD-L1 and TIM-3 blockade were both applied at day 0 (Figure 3B). Absolute antigen-specific counts were also increased in the DDB relative to the control condition (significant on days 21 and 28) (Figure 3C). Finally, DDB did not impact T-cell differentiation or exhaustion marker expression (Figure 3D-E). Independent cultures using isotype control antibodies confirmed the specific effects of anti-PD-L1 and anti-TIM-3 antibodies on the antigen-specific T-cell expansion (Supplementary Figure 1). We conclude that combined blockade of the PD-L1/PD-1 axis and TIM-3 can be incorporated in *ex vivo* cultures to increase antigen-specific CD8^+^ T-cell yield for adoptive immunotherapy without altering T-cell phenotypes.

**Figure 3.**
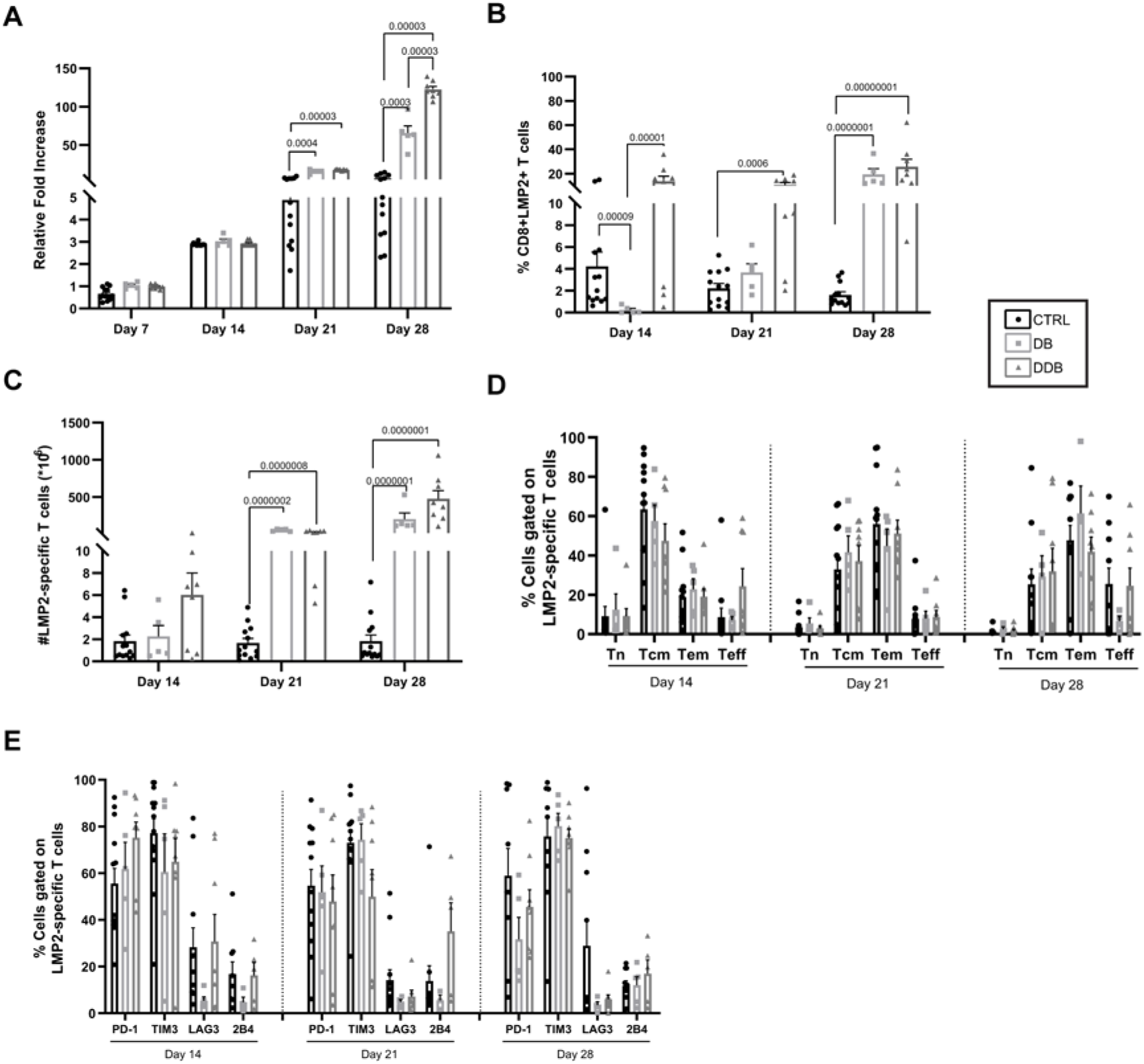
Delayed double blockade (DDB) further improves T-cell yield. (A) Total T-cell expansion relative to input at day 0 in the function of time and culture condition; no blocking antibodies (Ctrl), double anti-PD-L1 and anti-TIM-3 applied at culture initiation (double blockade – DB), anti-PD-L1 introduced at day 0 and anti-TIM-3 introduced at day 7 (delayed double blockade – DDB). (B) Percentages and (C) absolute counts of HLA-A0201-LMP2_426-434_ (LMP2) multimer positive T cells from the same cultures. (D) T-cell differentiation phenotypes and (E) immune checkpoint expression of LMP2 multimer positive cells in the same culture conditions and time-points (n=8 different donors). Significant (<0.05) p-values are indicated on the figure panels. All error bars represent SEM.

### Double immune checkpoint blockade generates functional antigen-specific T cells

Dual PD-L1 and TIM-3 blockade increases antigen-specific CD8+ T-cell expansion in culture without altering T-cell phenotypes suggesting comparable functionality. Intracellular cytokine secretion measurements and ELISpot assays at day 28 confirmed that a higher proportion of T cells were reactive upon LMP2_426-434_ peptide re-exposure in the DDB condition relative to control (no checkpoint inhibition) (Figure 4A-B). This was also generally the case when DDB was compared to double immune checkpoint blockade administered at day 0 (statistically significant in ELISPOT data at day 28). We thus pursued our functional assessments comparing DDB with the control condition. As further indication of increased functionality, a greater fraction of antigen-specific T cells expanded in the DDB condition relative to control expressed the proliferation marker Ki-67 and upon peptide re-exposure, more DDB exposed T cells displayed evidence of cytotoxic potential (surface CD107a and intracellular granzyme B expression) (Figure 4C-D). This was corroborated with cytotoxicity assays showing that T cells from the DDB condition were highly effective, especially at low effector:target ratios (Figure 4E-F). Importantly, in all our assays, combined PD-L1 and TIM-3 blockade did not lead to increased non-specific (off-target or spontaneous) cytokine release or cytotoxicity. Thus, dual immune checkpoint inhibition expands T-cell products with specific and augmented antigen reactivity.

**Figure 4.**
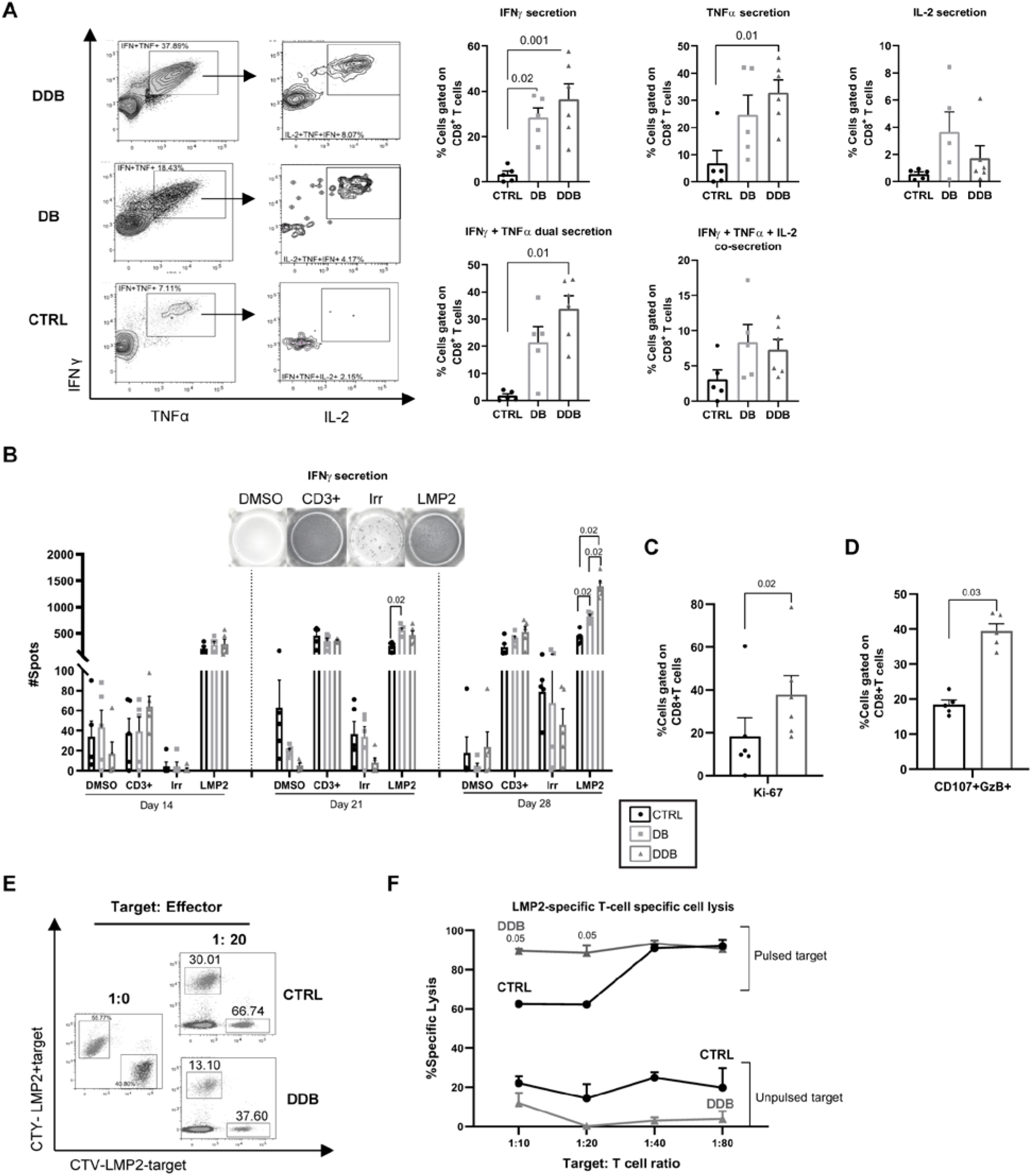
DDB expands a high proportion and number of functional antigen-specific T cells. (A) Representative dot plots showing intracellular cytokine secretion following LMP2_426-434_ antigenic restimulation at day 28 and compiled results from 5-6 independent donor cultures demonstrating the percentage of CD8^+^ T cells secreting IFNγ, TNFα, IL-2 and multiple cytokines; expansion with no blocking antibodies (CTRL), anti-PD-L1 and anti-TIM3 antibodies both introduced at the beginning of the culture (DB) or both antibodies but anti-TIM3 introduced at day 7 (DDB). (B) IFNγ ELISpot results using 50,000 cells per condition harvested from the cultures at the indicated time-points and using the following stimulating conditions: vehicle only (DMSO), anti-CD3 (CD3, positive control), irrelevant peptides (Irr.), LMP2_426-434_ peptide (LMP2). (C) Percentage of Ki-67 staining at day 28 among CD8^+^ T cells from the ctrl vs DDB cultures (n=5 different donors) and (D) co-expression of surface CD107a and intracellular granzyme-b (GzB) as surrogate for degranulation following LMP2_426-434_ exposure. (E) Fluorescence-based cytotoxicity assay (cell tracer yellow – CTY or cell tracer violet – CTV) showing loss of targets loaded (LMP2+) or not (LMP2-) with the LMP2_426-434_ antigen (numbers indicate percentages of total events) and (F) compiled results at different target:T-cell ratio (from 3 different donors). The significant p-values are indicated on the panels and error bars indicate SEM.

### PD-L1/TIM-3 blockade imparts no consistent gene expression signatures to expanded antigen-specific CD8^**+**^ T cells

To gain more insight into the biological impacts of double immune checkpoint blockade on antigen-reactive T cells, we submitted sorted day 28 multimer-positive T cells from three donors to paired transcriptome and TCR alpha-beta single-cell RNA sequencing (scRNA-Seq). Gene expression was compared between donors, conditions (no checkpoint blockade vs. DDB) and clonotypes. Global gene expression patterns across the three donors revealed strong donor-specific clustering (Figure 5A). The impact of DDB on gene expression for each donor antigen-specific T cells, was assessed using published human T-cell gene sets^17^. This enabled a detailed assessment of T-cell activation, proliferation, terminal differentiation, exhaustion and metabolism-associated gene expression as these processes are likely to be impacted by DDB. Overall, when all T cells from each condition were compared, no consistent change of gene expression signature could be identified across all donors (Figure 5B). We next evaluated whether clonotype-specific signatures could be identified. VD(J) sequencing revealed that the LMP2_426-434_ specific T cells were oligoclonal in all donors and conditions (1-4 clones representing more than 80% of all cells – Figure 5C and Supplementary Table 3). Most dominant clonotypes were shared between experimental conditions (albeit in some instances at different frequencies) and a few clonotypes were shared between donors (e.g. clonotype 1 and 3 common to donors 1 and 2, Supplementary Table 3). We next analyzed the expression of several pathways in the abundant clonotypes, defined as those identified in at least 10 cells and representing at least 1% of the repertoire in both the DDB and control conditions from the same donor. Similar to comparisons involving all clonotypes, no consistent pattern was found across donors when comparing the DDB versus control condition on a per clonotype basis (Figure 5D). Interestingly, our data unveiled divergent gene expression patterns for one clonotype (Clonotype 3) which was shared by Donor 1 and 2. Relative to its counterpart in the control condition, Donor 1 Clonotype 3 in the DDB condition expressed higher levels of genes related to T-cell exhaustion/differentiation and had a lowered expression of genes associated with T-cell proliferation. The same clonotype in the DDB condition from Donor 2 had, on the contrary, an increased expression of the genes associated with T-cell activation without any transcriptional changes related to T-cell dysfunction (T-cell exhaustion, terminal differentiation, low proliferation). Although obtained from a limited number of cells and donors, these results suggest that donor rather than clonotype related features may determine T-cell outcomes following DDB.

**Figure 5.**
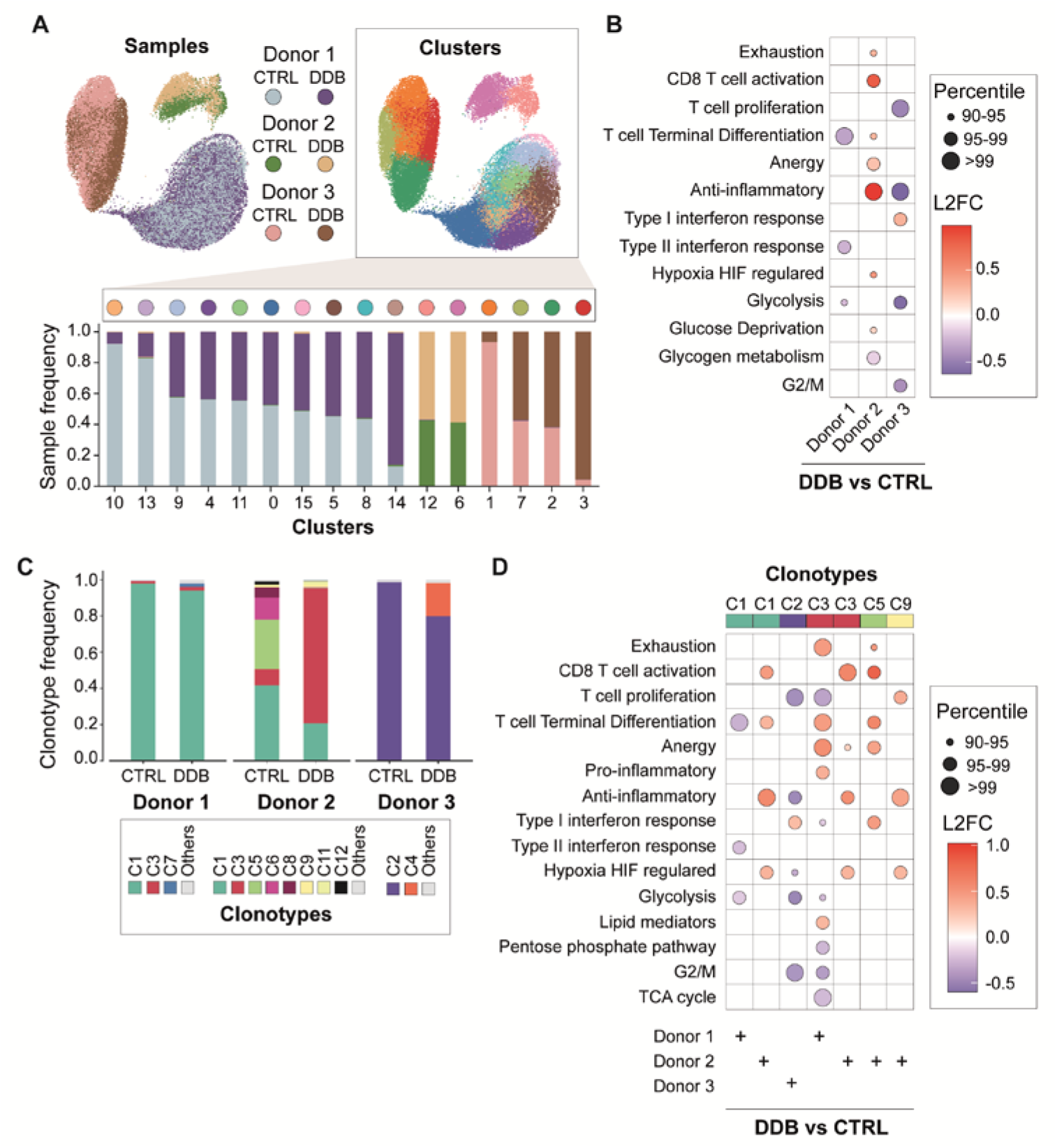
Single-cell RNA-sequencing of antigen-specific T cells after double immune checkpoint inhibition. (A) T-stochastic neighbor embedding (t-SNE) of normalized single-cell gene expression after dimensionality reduction from control (CTRL) and delayed double blockade (DDB) conditions, color coded by donor and experimental condition on the left and by cluster on the right. The barplot represents the percentage of each sample (colored as above left) in each cluster labeled on the x-axis and color-coded at the top of the graph. (B) Dot plot representing the change in the expression of genes related to pathways of interest in the DDB condition compared to the CTRL condition. The color of the dots represents the log2 fold-change of the genes in the pathway and the size of the dot is representative of the percentile ranking of the comparison in random gene sets (see Methods). (C) Clonotype frequencies in the CTRL and DDB conditions for each donor. (D) Similar to (B) comparing the cells of selected clonotypes/patients between the two experimental conditions.

To assess whether clonotype-specific transcriptional signatures may impact their expansion and clonal hierarchy within the cultures, we used bulk mRNA collected from sorted multimer negative and positive CD8+ T cells at day 21 and 28 from the same donors. RNA of suitable quality was obtained for 23 out of a potential of 24 samples (the multimer-positive fraction of the control condition from Donor 3 had to be excluded) and subjected to complementary determining region 3 (CDR3) region sequencing of the T-cell receptor-beta chain (TCRβ). Day 28 multimer-positive T-cell CDR3 sequencing matched very well paired VDJ sequencing of single cells (Supplementary Figure 2), confirmed the oligoclonality of the multimer-positive T cells at both day 21 and 28 and revealed that TCR repertoire diversity in the multimer-negative T cells was not affected by DDB (Figure 6A). Further, the multimer positive and negative fractions had limited overlap (Supplementary Figure 3), suggesting that sorting effectively separated most CD8+ antigen-specific cells from the rest of the T cells in culture. Clonotype hierarchy among multimer-positive T cells at day 21 and day 28 showed no consistent pattern of evolution in DDB relative to the control condition (Figure 6B-C). However, the proportion of certain clonotypes in these fractions (both DDB and control) varied importantly (>20%) between day 21 and 28. This was notably the case for Clonotype 3 from Donor 1 in the DDB condition, which declined markedly from day 21 to day 28, whereas this clonotype’s frequency changed only slightly between day 21 to 28 of DDB exposure in Donor 2 (Figure 6B). We then explored whether the clonotypes with altered abundance between day 21 and day 28 in the same donor and condition displayed a specific transcriptional profile at day 28. In the DDB condition, clonotype 3 of Donor 1, which showed a decreased abundance from day 21 to day 28, had a higher expression of genes related to T-cell activation, differentiation and exhaustion relative to other clonotypes from the same donor in the same condition (Figure 6D). Although less striking, we also noticed a weak trend of increased expression of genes related to exhaustion in clonotypes with decreased abundance at day 28 and a rise in the expression of genes related to proliferation in clonotypes with an increased abundance at day 28, irrespective of experimental conditions (Figure 6D). Taken together these data show that DDB has limited impact on the clonal diversity of the expanded T cells overtime. The proportion of the various clonotypes in time can nonetheless fluctuate in both culture conditions and that the gene expression signatures at day 28 offers possible explanations for such fluctuations. Globally, these results suggest that DDB confers no consistent transcriptional features to expanded antigen-specific CD8^+^ antigen-specific T cells but may alter activation/dysfunction and cell cycle-related processes in a clonotypes and donor-dependent fashion.

**Figure 6.**
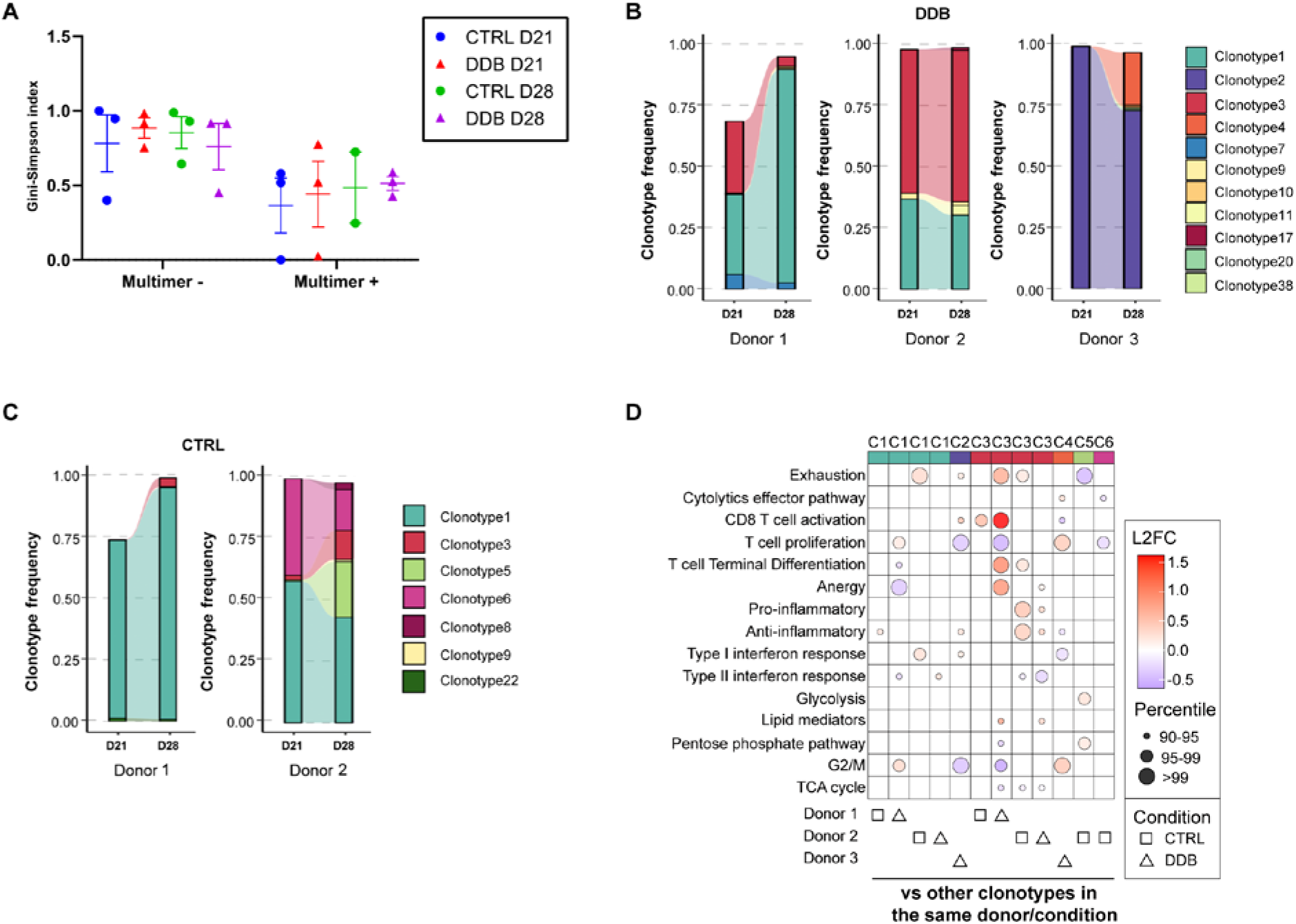
Impact of DDB on clonal diversity and stability in time. (A) Estimate of TCR repertoire diversity using the Gini-Simpson index among HLA-multimer negative and positive T cells at day 21 and 28. (B) Clonal hierarchy and clonal relatedness among HLA-A0201-LMP2_426-434_ multimer positive T cells between day 21 and 28 using the day 28 repertoire as reference in the DDB (B) or control condition (C) as determined by bulk CDR3 sequencing. The control condition in Donor 3 was not assessed due poor RNA quality at day 28. (D) Dot plot representing the change in the expression of genes related to pathways of interest when comparing a clonotype of interest in a donor/experimental condition to all the other clonotypes in the same donor/condition. The clonotypes selected are those whose abundance vary by more than 20% between day 21 and 28.

### The benefits of dual PD-L1 and TIM-3 blockade extend to TAA-specific T cells

The EBV-derived LMP2_426-434_ antigen stimulates a memory T-cell repertoire in more than 90% of adults^23^. The *ex vivo* expansion of naïve T cells is generally considered more challenging for several reasons such as size of the repertoire and amount of stimulation required. We sought to determine the impact of dual PD-L1/TIM-3 blockade on the priming and expansion of naïve T cells. It was previously shown that CD8+ T cells specific against Wilm’s tumor 1 (WT1) derived peptides, a clinically relevant tumor-associated antigen (TAA), are phenotypically naïve in healthy individuals^24^ but can be expanded from a majority of such donors^25 26^. Volunteer donor T cells were stimulated with an HLA-0201 restricted WT1 peptide (WT1_37-45_) using the same stimulation/expansion conditions for LMP2_426-434_ specific T cells. WT1-specific T cells expanded following a similar pattern as LMP2_426-434_-specific T cells (Figure 7A-C). The expansion, percentages and number of antigen-specific cells as well as their cytokine secretion were improved in the DDB condition relative to cultures without immune checkpoint blockade, again without altering T-cell differentiation patterns or exhaustion marker expression (Figure 7D-H). We conclude that our results obtained with LMP2_426-434_ extend beyond virus-specific memory T-cell expansion and that double PD-L1/TIM-3 immune checkpoint blockade can improve the expansion of functional TAA-specific CD8+ T cells from naïve repertoires.

**Figure 7.**
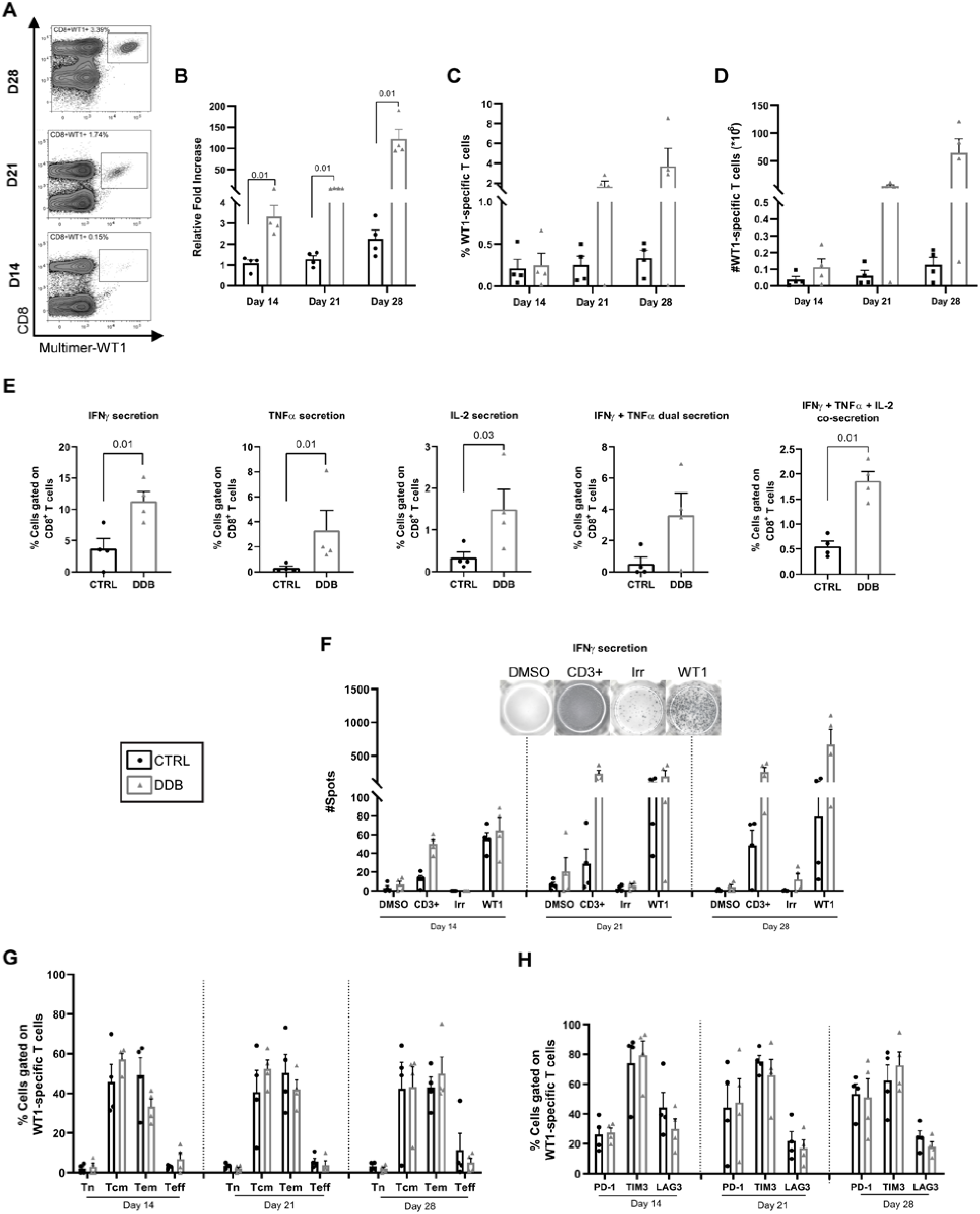
Improved expansion of functional WT1-specific T cells with DDB. (A) Representative HLA-A0201-WT1_37-45_ multimer staining (WT1) of CD8^+^ T cells. (B) Cell expansion expressed as fold increase relative to cell input at the beginning of the culture (15×10^6^) in ctrl (no blocking antibodies) or delayed double anti-PD-L1 and anti-TIM3 blockade (DDB) condition, as well as percentage (C) and absolute numbers (D) of WT1-specific T cells at different time-points in culture. (E) Proportion of intracellular cytokine expression (IFNγ, TNFα, IL-2, and multiple cytokines) following WT1 exposure at day 28 of the culture in CD8^+^ cells and (F) IFN-γ ELISpot results at days 14, 21, and 28 of culture following exposure to vehicle alone (DMSO), peptides not used in the culture (irrelevant, Irr) as negative controls, anti-CD3ε (positive control) and the targeted peptide (WT1). (G) T-cell differentiation; naïve (Tn), central memory (Tcm), effector memory (Tem), and effector T cells (Teff) and (H) percentage of immune checkpoint surface expression on WT-1 specific T cells. Four (4) different donors, p-values are indicated on the figures, error bars indicate SEM.

## Discussion

The stimulation and expansion of antigen-specific T cells for adoptive immunotherapy is an attractive strategy to target a wide variety of cancer antigens. However, T-cell expansion is limited by the expression of inhibitory receptors and the development of T-cell dysfunction. We confirmed and extended previous studies revealing that immune checkpoint receptors are expressed on repeatedly activated T cells and that their corresponding ligands can be present on antigen-presenting cells used *ex vivo*. Co-expression of PD-1 and TIM-3 has been used to describe highly exhausted CD8^+^ T cells, and combined blockade of these receptors in murine models has previously been shown to improve tumor control relative to single blockade^19 20 27^. We show that antibody-mediated immune checkpoint blockade targeting PD-1 or TIM-3 alone is insufficient to improve CD8^+^ T-cell expansion while the combination improves T-cell expansion and antigen-specific reactivity. These data confirm the synergistic potential of immune checkpoint blockade and the particular relevance of TIM-3 and PD-1 as inhibitory receptors in CD8^+^ T cells^20^. The reasons for this synergistic effect might be due to the mobilization of different signaling intermediates by the two receptors. While PD-1 relies on the recruitment of SHP phosphatases to mediate its effects (as for several other negative co-stimulatory molecules), TIM-3 uses Bat3 and Fyn for its stimulatory and inhibitory effects, respectively. We reasoned that delaying TIM-3 blockade by one week after the start of the culture even in the presence of TIM-3 ligands on APC would be beneficial, given previous experimental evidence that TIM-3 provides activation signals following a first T-cell stimulation^28 29^. This contrasts with PD-1, which conveys inhibitory signals in both exhausted T cells and early after activation^30^, justifying the use of anti-PD-L1 at culture initiation. We realize that several refinements may be required to fully leverage the potential agonistic/antagonistic effects of TIM-3 on T-cell activation and expansion and determine whether anti PD-L1/PD-1 blockade may also be improved by altering the timing of the inhibition. In addition, whether dual PD-L1/TIM-3 blockade improves T-cell expansion and function across multiple types of T-cell manufacturing protocols should be tested, just like further combinations with blockade of additional inhibitory receptors. Nonetheless, our work provides a new strategy for T-cell expansion and offers novel biological insights on the effects of PD-L1/PD-1 axis and TIM-3 blockade in human antigen-specific T-cell clonotypes submitted to multiple antigenic stimulations.

Our data support that sustained dual PD-L1/TIM-3 blockade through several rounds of antigenic stimulations provides ongoing benefits without exacerbating T-cell dysfunction or curbing expansion as inferred by previous studies of using inhibitory receptor gene deletion^31^. This was assessed through phenotyping, functional assays, and gene expression in single cells. Our results indicate that antigen-specific CD8^+^ T cells expanded under combined TIM-3/PD-L1 blockade are functional as well as specific and that their clonal composition is generally stable in time and relative to their counterpart not exposed to immune checkpoint blockade. It has been reported that immune checkpoint blockade *in vivo* can shape the T-cell repertoire^32-34^. Thus, from a perspective of T-cell therapy, it was essential to define whether an intervention during *ex vivo* T-cell expansion alters the clonal identity of the cellular product. While we did not observe consistent effects of PD-L1/TIM-3 blockade on T-cell transcriptome or clonotype distribution, altered transcriptional profiles in a clonotype and donor-dependent manner nonetheless suggests that immune checkpoint blockade in adoptive T-cell therapy may require personalization.

Further work will be required to understand why some clonotypes may be more susceptible to differentiation/exhaustion following immune checkpoint blockade at different time-points (previous activation/proliferation history, differentiation status at the beginning of checkpoint blockade, etc). Nevertheless, our data suggest that even after 28 days in culture, the antigen-specific T cells are highly functional, with a third expressing Tcm markers. This indicates that antigen-specific T cells generated in high numbers after dual immune checkpoint blockade could further expand after transfer and likely respond to further immune checkpoint blockade administered *in vivo*.

We conclude that dual PD-L1/TIM-3 blockade is a readily applicable strategy to improve the expansion of functional antigen-specific CD8^+^ T cells expansion *ex vivo* for adoptive immunotherapy.

## Data Availability

The data can be found in the Gene Expression Omnibus (GSE182537 and GSE181682).

## Acknowledgments

The authors are grateful to the volunteer blood donors and Héma-Québec for the leukocytes reduction chamber procurement and handling. We also acknowledge the valuable contribution of Martine Dupuis (flow cytometry and sorting) and the Genome Québec staff for the single-cell RNA-sequencing. SL and VJ are respectively former Cole Foundation and Fonds de recherche du Québec-Santé (FRQS) studentship awardees, JSD and VPL hold FRQS clinician-scientist career awards and JSD is a member of the ThéCell network and of the Canadian Donation and Transplant Research Program (CDTRP).

## List of Abbreviations

ACT: Adoptive Cell Therapy
APC: Antigen Presenting Cell
CCR7: C-C chemokine receptor type 7
CD: Cluster of differentiation
CDR: Complementarity Determining Region
CTLA-4: Cytotoxic T-lymphocyte-associated protein 4
CTLs: Cytotoxic T-lymphocytes
CTV: Cell Trace Violet
CTY: Cell Trace Yellow
DC: Dendritic Cell
DMSO: Dimethylsulfoxide
EBV: Epstein-Barr virus
ELISPOT: Enzyme-linked immune absorbent spot
FBS: Fetal Bovine Serum
FOXP3: Forkhead Box P3
Gal-9: Galectin-9
GM-CSF: Granulocyte-Macrophage Colony-Stimulating Factor
GrzB: GranzymeB
HLA: Human Leukocyte Antigen
IFN: Interferon
IL: Interleukin
KLRG1: Killer cell Lectin-like Receptor G1
LAG-3: Lymphocyte-Activation Gene 3
LRSC: Leukocyte Reduction System Chamber
mAB: Monoclonal Antibody
MHC: Major Histocompatibility Complex
NGS: Next-generation Sequencing
PBMC: Peripheral Blood Mononuclear Cell
PBS: Phosphate-buffered saline
PD-1: Programmed Cell Death-1
PD-L1: Programmed Death Ligand-1
PGE2: Prostaglandin E2
PHA: T-cell mitogen phytohemagglutinin
PMA: Phorbol 12-myristate 13-acetate
RT: Room Temperature
sc-RNA: Single Cell RNA
TAA: Tumor-Associated Antigen
Tcm: Central Memory T cell
TCR: T-cell Receptor
Teff: Effector T cell
Tem: Effector Memory T cell
TIM-3: T-cell Immunoglobulin and Mucin-domain Containing-3
TMB: 3,3′,5,5′-Tetramethylbenzidine
TNF: Tumor Necrosis Factor
TSA: Tumor-Specific Antigen
WT1: Wilms’ tumor suppressor gene1

