## Supplementary Information for "Combined PD-L1 and TIM-3 blockade improves the expansion of fit human CD8+ antigen-specific T cells for adoptive immunotherapy"

* Jean-Sébastien Delisle.

**This PDF file includes:**

Tables S1 to S3
Figures S1 to S3

**Supplementary Table S1:** Single cell filtering results. Initial number of barcodes represents the output from CellRanger v3.0.2 while the number of cells after filtering corresponds to the cells kept for downstream analysis.

| Sample | Initial number of barcodes | Number of cells after filtering |
|---|---|---|
| CTRL1 | 20182 | 12753 |
| CTRL2 | 8550 | 1950 |
| CTRL3 | 27019 | 9109 |
| DB1 | 19724 | 10101 |
| DB2 | 11107 | 2665 |
| DB3 | 30813 | 8981 |

**Supplementary Table S2:** Gene list used for analysis.

| TCA cycle | G2-M | Pentose Phosphate Pathway | | Lipid mediators | | Glycogen Metabolism | Glycogen Metabolism | Glucose Deprivation |
|---|---|---|---|---|---|---|---|---|
| ACLY | AURKA | G6PD | ADIPOQ | GH1 | FASN | GBE1 | GBE1 | NRN1 |
| ACO1 | BIRC5 | G6PDH | TNFRSF9 | HGF | IRS2 | GYS1 | GYS1 | FAM129A |
| ACO2 | BUB1 | PGLS | AgRP | ICAM1 | KLF15 | GYS2 | GYS2 | IL23A |
| CS | BUB1B | PRPS1 | ANGPT1 | IGFBP1 | PPARG | UGP2 | UGP2 | SPRY1 |
| DLAT | CCNA2 | PRPS1L1 | ANGPT2 | IGFBP2 | PPARGC1A | AGL | AGL | GADD45B |
| DLD | CCNB1 | PRPS2 | ANGPTL3 | IGFBP3 | RETN | PGM1 | PGM1 | HSPA1A |
| DLST | CCNB2 | RBKS | ANGPTL4 | IGFBP4 | SIRT3 | PGM2 | PGM2 | HSPA1B |
| FH | CCNF | RPE | ANGPTL6 | IGFBP6 | SLC2A4 | PGM3 | PGM3 | GPNMB |
| IDH1 | CDC20 | RPIA | TNFSF13B | IGFBP7 | SREBF1 | PYGL | PYGL | HAVCR2 |
| IDH2 | CDC25B | TALDO1 | BMP4 | IL1b | ACACB | PYGM | PYGM | SOAT2 |
| IDH3A | CDC25C | TKT | CTSD | IL6 | AXIN1 | GSK3A | GSK3A | ADAMTS6 |
| IDH3B | CDK1 | | CTSS | IL10 | CCND1 | GSK3B | GSK3B | |
| IDH3G | CDKN2D | | CCL2 | IL11 | CDK4 | PHKA1 | PHKA1 | |
| MDH1 | CENPA | | CCL5 | INS | CEBPB | PHKB | PHKB | |
| MDH1B | CENPF | | RARRES2 | LEP | CEBPD | PHKG1 | PHKG1 | |
| MDH2 | CKS2 | | CFD | LIF | DKK1 | PHKG2 | PHKG2 | |
| OGDH | KIF20A | | CRP | LCN2 | E2F1 | | | |
| PC | PLK1 | | CXCL8 | CSF1 | FABP4 | | | |
| PCK1 | RACGAP1 | | DPP4 | MIF | FASN | | | |
| PCK2 | TOP2A | | ESM1 | MPO | | | | |
| PDHA1 | | | S100A12 | GHRL | | | | |
| PDHB | | | AHSG | GHSR | | | | |
| SDHA | | | FGF1 | LIPE | | | | |
| SDHB | | | FGF2 | LPL | | | | |
| SDHD | | | FGF21 | AGT | | | | |
| SUCLA2 | | | FGF23 | CEBPA | | | | |
| SUCLG1 | | | LGALS3 | CFD | | | | |
| SUCLG2 | | | GCG | FABP4 | | | | |

**Supplementary Table S2:** Gene list used for analysis (continued).

| Glycolysis | Hypoxia-HIF regulated | | Type II Interferon Response | Type I Interferon Response | Anti-inflammatory | Pro-inflammatory | Anergy | T- Cell Terminal Differentiation |
|---|---|---|---|---|---|---|---|---|
| ALDOA | LAMB1 | IL10RA | IFNG | IRF1 | TIGIT | IL1A | EGR3 | TIGIT |
| ALDOB | ALDOA | IL15RA | CXCL9 | IFIH1 | IDO1 | IL1B | NR4A3 | PDCD1 |
| ALDOC | ADM | ITGA6 | CXCL10 | IFITM3 | LGALS3 | TNF | EGR1 | CD274 |
| BPGM | BCL2 | ITK | STAT1 | DDX58 | PDCD1 | IFNG | NR4A2 | CTLA4 |
| ENO1 | BCL2L1 | JUND | CD274 | IFI44L | FOXP3 | TBX21 | EGR2 | LAG3 |
| ENO2 | FOS | IGHG4 | CDKN1A | IFI6 | ENTPD1 | CCL3 | TNFSF11 | HAVCR2 |
| GALM | JUN | MAP3K5 | MYC | IFITM2 | CD274 | CCL4 | IRF4 | CD244 |
| GCK | SRC | MAP2K1 | SMAD7 | NAMPT | CSF2 | PRF1 | GCH1 | CD160 |
| GPI | CREBBP | MAP2K2 | IRF1 | OASL | CTLA4 | GZMA | GADD45B | |
| HK2 | CCR6 | MIF | HLA-F | RTP4 | CXCL12 | GZMB | NFATC1 | |
| HK3 | CEBPB | NFATC1 | HLA-G | TREX1 | CXCL5 | GZMK | JARID2 | |
| PFKL | ENO1 | NFKB2 | HLA-A | ADAR | CXCL8 | GZMH | SLC29A3 | |
| PGAM2 | FAS | NFKBIE | HLA-E | TENT5C | MIF | CD8A | HLF | |
| PGK1 | FASLG | TP53 | HLA-C | LY6E | PTGS2 | FASLG | ZFP36L1 | |
| PGK2 | FKBP4 | TGM6 | HLA-B | MCOLN2 | VEGFA | CCL2 | RNF19A | |
| PGM1 | GAPDH | TNF | HLA-DRA | APOBEC3G | | CCL20 | ADORA2B | |
| PGM2 | SLC2A1 | TNFRSF1B | HLA-DRB5 | IL15 | | IL2 | NOCT | |
| PGM3 | SLC2A3 | TRADD | HLA-DRB1 | ISG15 | | IL6 | DUSP6 | |
| PKLR | CSF2 | TNFSF10 | HLA-DQA1 | MX1 | | IL12a | NDRG1 | |
| TPI1 | IFNG | TRAP1 | HLA-DQB1 | TLR3 | | IL17a | ADORA2A | |
| | IFNB1 | VEGFA | HLA-DQB1-AS1 | | | IL23a | HSPA1A | |
| | IL13 | | HLA-DQA2 | | | PTGS2 | PFKP | |
| | IL1R1 | | HLA-DQB2 | | | TLR4 | FYN | |
| | IL2RA | | HLA-DOB | | | TNF | CTSE | |
| | IL4 | | HLA-DMB | | | | ZNF629 | |
| | IL5 | | HLA-DMA | | | | F2R | |
| | IL6R | | HLA-DOA | | | | RNF128 | |
| | CXCL8 | | HLA-DPA1 | | | | LAG3 | |
| | | | HLA-DPB1 | | | | RGS16 | |
| | | | HLA-DPA1 | | | | E2F1 | |
| | | | | | | | E2F2 | |

**Supplementary Table S2:** Gene list used for analysis (continued).

| T-Cell Proliferation | CD8 T-Cell Activation | Cytolytics effector pathway | Exhaustion |
|---|---|---|---|
| UBE2T | CD69 | EOMES | TOX |
| CENPU | CCR7 | TBX21 | TOX2 |
| CHEK1 | CD27 | GZMB | EOMES |
| AURKA | BTLA | PRF1 | TBX21 |
| CAPG | CD40LG | FASLG | NR4A2 |
| PCLAF | IL2RA | GZMH | PRDM1 |
| BUB1B | CD3E | GZMA | PDCD1 |
| CCNB2 | CD47 | | HAVCR2 |
| DLC1 | EOMES | | CD244 |
| CDK1 | GNLY | | CD160 |
| CCNA2 | GZMA | | LAG3 |
| SERPINB2 | GZMB | | CD38 |
| PCNA | PRF1 | | ENTPD1 |
| TOP2A | IFNG | | CD101 |
| STAT5A | CD8A | | IFNG |
| Il2 | CD8B | | TNF |
| | FASLG | | IL2 |
| | LAMP1 | | CTLA4 |
| | LAG3 | | |
| | CTLA4 | | |
| | HLA-DRA | | |
| | TNFRSF4 | | |
| | ICOS | | |
| | TNFRSF9 | | |
| | TNFRSF18 | | |

**Supplementary Table S3:** TCR alpha/beta chain sequences of the most abundant clonotypes across donors and conditions.

| Clonotype ID | nt_TRA | nt_TRB | aa_TRA | aa_TRB | Donor |
|---|---|---|---|---|---|
| clonotype1 | TGTGCTGCCCTCATGGATAGCAACTATCAGTTAATCTGG | TGCGCCAGCAGTGATGACGGGATGAACACTGAAGCTTTCTTT | CAALMDSNYQLIW | CASSDDGMNTEAFF | 1 and 2 |
| clonotype2 | TGTGCTGTGCTCATGGATAGCAACTATCAGTTAATCTGG | TGCGCCAGCAGTGGGGACGGTATGAACACTGAAGCTTTCTTT | CAVLMDSNYQLIW | CASSGDGMNTEAFF | 3 |
| clonotype3 | TGTGCAATGAGCGCGGAGGAGGACATGCGCTTT | TGTGCCAGCAGCTCCCGGCTAGCGGGGATCCAAACGAGCTCCTACAATGAGCAGTTCTTC | CAMSAEEDMRF | CASSSRLAGIQTSSYNEQFF | 1 and 2 |
| clonotype4 | TGTGCAATGAGCGCGGAATCAAATTCCGGGTATGCACTCAACTTC | TGTGCCAGCAGCTTAGGGGGTTATGAGCAGTTCTTC | CAMSAESNSGYALNF | CASSLGGYEQFF | 3 |
| clonotype5 | TGTGCAGGGGCCGGGGCTGGGAGTTACCAACTCACTTTC | TGCGCCAGCAGCTTGGAGGGACAGGCGAGCTCCTACGAGCAGTACTTC | CAGAGAGSYQLTF | CASSLEGQASSYEQYF | 2 |
| clonotype6 | TGTGCTGTGACTAACTTTGGAAATGAGAAATTAACCTTT | TGCGCCAGCAGCCAACTACAGGGATACGAGCAGTACTTC | CAVTNFGNEKLTF | CASSQLQGYEQYF | 2 |
| clonotype7 | TGTGCAATGAGAGAGCCTCTCACGGGAGGAGGAAACAAACTCACCTTT | TGTGCCAGCAGTGGACTAGCGATCTCCTACGAGCAGTACTTC | CAMREPLTGGGNKLTF | CASSGLAISYEQYF | 1 |

nt: nucleotide; aa: amino acid; TRA: TCR alpha chain; TRB: TCR beta chain.

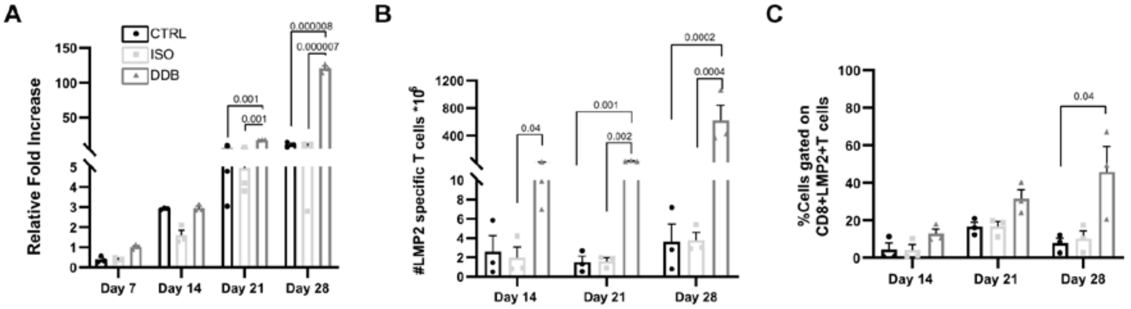


**Supplementary Figure S1. Validation of the DDB strategy using isotype control antibodies**. (A) Cell expansion relative to input at the beginning of the culture ($15x10^6$) in function of time and culture condition; no antibodies (ctrl), isotypes (ISO) according to the Delayed double blockade (DDB) scheme or DDB. Absolute count (B) and percentage (C) of HLA-A0201-LMP2$_{426\text{-}434}$ (LMP2) multimer positive T cells in the same conditions and at the same time-points. 3 different donors, significant p-values are indicated, error bars indicate SEM.

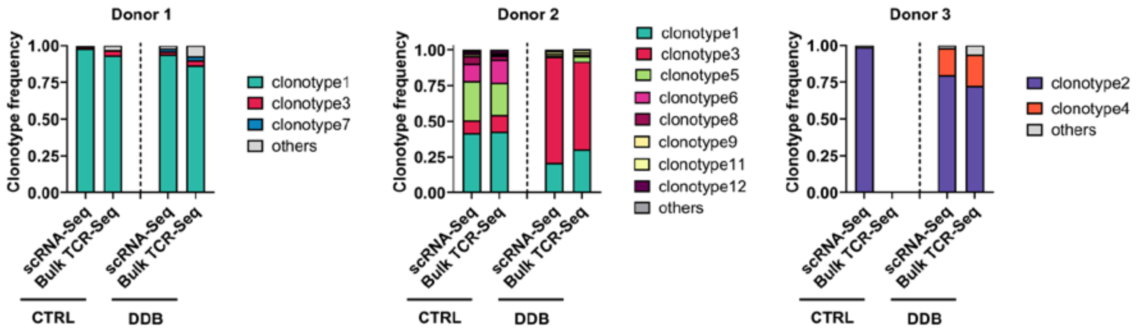


**Supplementary Figure S2. Correlation between clonotype determination using single-cell RNA sequencing (scRNA-seq) and bulk CDR3 beta-chain TCR sequencing**. Comparison of clonotype repertoire among sorted HLA-A0201-LMP2$_{426-434}$ multimer positive T cells at day 28 from the control condition (CTRL) and delayed double blockade (DDB) condition according to scRNA-seq or bulk CDR3 sequencing (bulk TCR-Seq) for each donor. The bulk TCR-Seq from the CTRL condition in Donor 3 was not considered due to poor RNA quality.

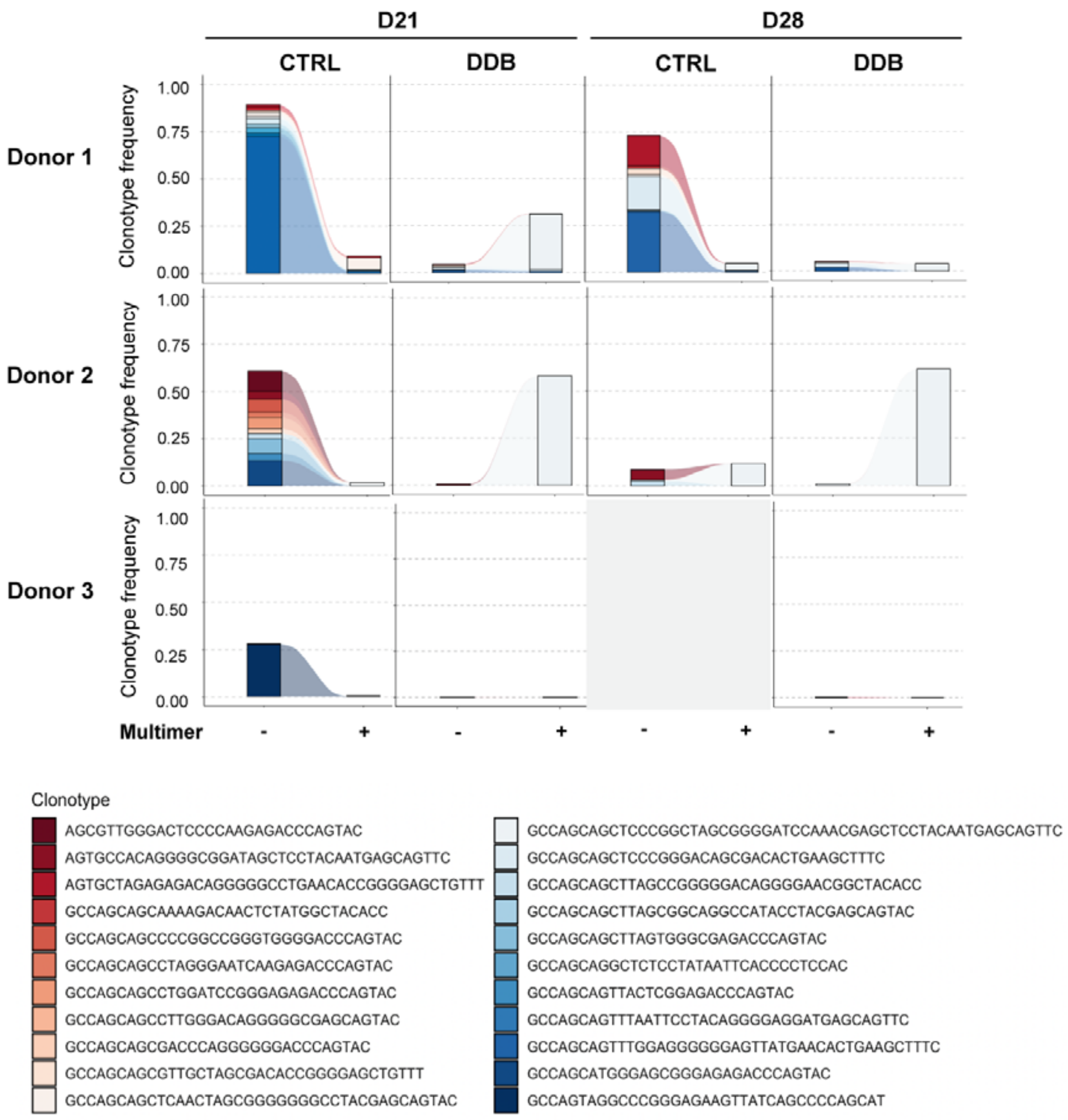


**Supplementary Figure S3. Limited overlap between HLA-A0201-LMP2$_{426\text{-}434}$ multimer positive and negative T cells.** Bulk CDR3 beta-chain TCR sequencing data from sorted multimer positive (+) and negative (-) fractions from all donors in both experimental condition (control - CTRL and delayed double blockade – DDB) at the day 21 and 28 time points showing limited clonotype overlap. Clonotypes present in both fractions (identified by beta-chain CDR3 sequences) are represented with none being abundant (>10%) in both fraction simultaneously. Due to poor RNA quality, the multimer-positive fraction of the control condition in Donor 3 could not be analyzed.
